# Beyond the reCOVer trial - nine-month functional outcomes after rovunaptabin (BC007) in post-COVID syndrome: a prospective cohort study

**DOI:** 10.64898/2026.11.24.26368572

**Authors:** Bettina Hohberger, Alexander Smit, Gerd Wallukat, Thomas Knauer, Maria Reppa, Ronja Zwerenz, Valerie Hartig, Merle Flecks, Stephanie Zellinger, Philipp Fleischmann, Martin Herrmann, Petra Lakatos, Cornelius Rosenkranz, Jakob Hoffmanns, Marion Ganslmayer, Friedrich Kruse, Stefanie Maas, Tobias Borst, Ralph Heimke-Brinck, Andreas Stog, Eva Rühl, Victoria Zeisberg, Adam Skornia, Alexander Bartsch, Armin Ströbel, Monika Wytopil, Caroline Merkel, Sophia Hofmann, Katja G. Schmidt, Julia Schottenhamml, Christian Mardin, Jürgen Rech, Thomas Harrer

**Author notes:** Corresponding author: PD Dr. Dr. Bettina Hohberger, Schwabachanlage 6, 91054 Erlangen.

## Abstract

**Background:** Long-term functional outcomes after exposure to rovunaptabin (BC007) in patients with post-COVID syndrome (PCS) still remain elusive.

**Methods:** We compared 24 participants previously exposed to rovunaptabin in the phase IIa reCOVer trial with 64 independently recruited participants of disCOVer study who had never received rovunaptabin. All had PCS with fatigue and functional autoantibodies against G-protein-coupled receptors. Median follow-up from baseline was 267.5 and 278 days, respectively. We compared follow-up Bell scores using analysis of covariance adjusted for baseline Bell score, age, and sex. Other patient-reported outcomes were summarized descriptively in the previously exposed cohort.

**Findings:** Mean Bell scores increased from 34.58 to 51.25 in the rovunaptabin exposed cohort and from 40.47 to 46.25 in the rovunaptabin-naive cohort. The adjusted difference in follow-up Bell score was 9.71 points in favor of the exposed cohort (95% CI 1.80-17.63, p = 0.017). This cohort showed an increased mean FACIT-Fatigue score by 9.71 points and the mean Fatigue Severity Scale score decreased by 1.02 points.

**Interpretation:** Prior one-time rovunaptabin exposure was associated with better self-reported functional status at approximately nine months after rovunaptabin exposure. Controlled studies with longer follow-up are needed to assess whether rovunaptabin produces lasting functional benefit, even when administered multiple times.

**Funding:** The study was funded by the Bavarian State Ministry of Science and Arts, the German Research Foundation, and the German Federal Ministry for Research, Technology and Space.

**Research in context:** *Evidence before this study:* Post-COVID syndrome (PCS), also known as Post-Acute Infection Syndrome (PAIS), post-acute sequelae of SARS-CoV-2 (PASC) or Long-COVID (LC), summarizes different clinical phenotypes with clinical symptoms longer than three months after acute COVID-19. As the specific molecular pathways for each PCS phenotype are still under investigation, regular patients’ care is based on a symptomatic, multidisciplinary approach. One of those PCS patients’ phenotypes is characterized by an autoimmune phenotype with a seropositivity of functional autoantibodies targeting G-protein-coupled receptors (GPCR-fAAbs). Preceding in-vitro and in-vivo data showed a neutralization of the GPCR-fAAb by the aptamer rovunaptabin (BC007). The reCOVer trial was the first prospective, randomized, double-blind, placebo-controlled phase IIa clinical trial investigating rovunaptabin in GPCR-fAAb seropositive patients with PCS. We searched PubMed from inception until November 23rd, 2026 using search terms “BC007”, “BC007”, “rovunaptabin”, “follow-up”, and “post-COVID” without language restrictions. This search identified 203 results, of which only two were considered relevant to the research question: a case report describing an individual treatment attempt in a single patient and the initial report of the reCOVer trial. A further phase II clinical trial (BLOC) investigating rovunaptabin in PCS patients was registered in the database clinicaltrials.gov (ID: NCT05911009), yet no study data have been published until now.

*Added value of this study:* This is the first study investigating functional outcomes nine months after administration of rovunaptabin in GPCR-fAAb seropositive patients with PCS. By comparing previously exposed patients from the phase IIa reCOVer clinical trial with an independently recruited cohort of GPCR-fAAb seropositive patients with PCS, who had never received rovunaptabin, this comparative approach showed an improvement of functional outcomes beyond the initial treatment period in the context of an independent, non-rovunaptabin exposed cohort nine months after one-time application.

*Implications of all the available evidence:* The findings provide additional evidence on the longer-term functional course of GPCR-fAAb seropositive patients with PCS after administration of rovunaptabin. Given the significant disease burden and continued lack of targeted therapeutic approaches, these results may help to design future clinical trials investigating rovunaptabin in PCS patients’ population.

## 1. Introduction

In late 2019, severe acute respiratory syndrome coronavirus 2 (SARS-CoV-2) emerged and led to the coronavirus disease 2019 (COVID-19) pandemic, with approximately 800 million reported cases and 7 million recorded deaths worldwide [1]. Acute illness ranges from mild to critical and may include fever, cough, and myalgia [2]. Some patients experience persistent symptoms for months after the acute infection, a condition referred to here as post-COVID syndrome (PCS). Its exact pathogenesis remains elusive. Persistent inflammation, endothelial dysfunction, viral persistence, microclot formation, and autoimmunity have been proposed as contributing mechanisms and may be relevant to different patient subgroups [3]. In a subset of patients, functional autoantibodies against G-protein-coupled receptors (GPCR-fAAbs) have been proposed to contribute to the clinical features of PCS [4]. These autoantibodies can modulate receptor-mediated signaling [5]. Previous studies detected functional autoantibodies (fAAbs) during acute COVID-19 and suggested that they might contribute to PCS [5]. Wallukat et al. also reported GPCR-fAAbs in sera from patients with PCS [6]. GPCR-fAAbs have been linked to several other conditions, including glaucoma, myocarditis, and chronic fatigue syndrome [7]. Investigational strategies aim to remove or neutralize these autoantibodies, inhibit their effects on receptor signaling, or reduce their production.

Aptamers are single-stranded oligonucleotides known to bind specific molecular targets with high affinity [8]. Rovunaptabin (BC007) is a synthetic DNA aptamer originally developed as a thrombin inhibitor [9]. Its development as an anticoagulant was discontinued because of its short duration of action. Wallukat et al. subsequently showed that rovunaptabin neutralized fAAbs against the β1-adrenergic receptor in samples from patients with dilated cardiomyopathy in vitro [6]. Subsequent studies reported neutralization of GPCR-fAAbs targeting the β2- and α1-adrenergic receptors, the endothelin type A receptor, and the angiotensin II type 1 receptor [10]. A three-part phase I study evaluated the safety, tolerability, and pharmacokinetics of rovunaptabin in healthy participants and reported that it was generally well tolerated [11]. An initial report of individual treatment and a subsequent randomized, double-blind, placebo-controlled phase IIa trial involving 29 patients with PCS reported GPCR-fAAb neutralization and improvements in selected fatigue and quality-of-life outcomes [12, 13].

Earlier studies provided limited information on longer-term outcomes. In this study, GPCR-fAAb seropositivity at baseline was used to define a seropositive PCS subgroup reflecting an autoimmune mechanism rather than to track whether autoantibodies disappear over time. The present study therefore focused on comparing longer-term symptom-related functional status approximately nine months after baseline - rather than differences in disease duration per se - between GPCR-fAAb-positive participants previously exposed to rovunaptabin in the reCOVer trial and an independently recruited cohort of GPCR-fAAb-positive participants, with PCS who had never received rovunaptabin.

## 2. Materials and Methods

### 2.1 Study design and participants

Participants were recruited from the patients’ population at the Department of Ophthalmology, Uniklinikum Erlangen, who had previously participated in the disCOVer study or the reCOVer clinical trial. Participants of reCOVer clinical trial were previously participants of the disCOVer study. The present analysis is a sub-analysis of the complete observational trial disCOVer trial, showing data of 93 GPCR-fAAb seropositive patients with PCS. Eligibility required age >18 years, a PCR-confirmed SARS-CoV-2 infection, PCS symptoms persisting for more than three months, including fatigue. Neither a specific disease duration nor a defined interval since the positive SARS-CoV-2 test served as an inclusion criterion or stratification factor for this follow-up comparison. A part of these patients was allocated to a secondary therapeutic trial reCOVer. The reCOVer trial (EudraCT 2022-001781-35) was a prospective, randomized, double-blind, placebo-controlled phase II clinical trial with crossover treatment at the Uniklinikum Erlangen, Germany. Its design and initial results have been published [13]. All 29 reCOVer participants received rovunaptabin during the trial. Follow-up analysis of further disCOVer participants, never received rovunaptabin, was done (n=64). Of the reCOVer participants, 24 participated in this follow-up and received no additional rovunaptabin after trial completion. Table 1 summarizes follow-up participation and baseline characteristics. Figure 1 shows the present trial.

**Table 1.** Demographic and baseline clinical characteristics, disease duration, and follow-up intervals of participants with post-COVID syndrome in the rovunaptabin-exposed (reCOVer) and rovunaptabin-naive (disCOVer) cohorts. Data are presented as mean (SD), median (range), or number of participants (n), as appropriate. Disease duration was approximated by the interval between the recorded positive SARS-CoV-2 test and baseline assessment.

| **Characteristic** | **reCOVer<br>previous rovunaptabin exposure participants<br>(n=24)** | **disCOVer<br>rovunaptabin-naive participants<br>(n=64)** |
|---|---|---|
| **Age at baseline, years, mean (SD)** | 39.33 (9.00) | 41.47 (11.76) |
| **Women/Men (n)** | 7/17 | 38/26 |
| **Baseline Bell score, mean (SD)** | 34.58 (12.85) | 40.47 (17.50) |
| **Baseline to follow-up, days, median (range)** | 267.5 (175-392) | 278 (150-417) |
| **Positive test to baseline, days, median (range)** | 1,142 (510-1,531) | 268.5 (87-673) |

**Figure 1.**
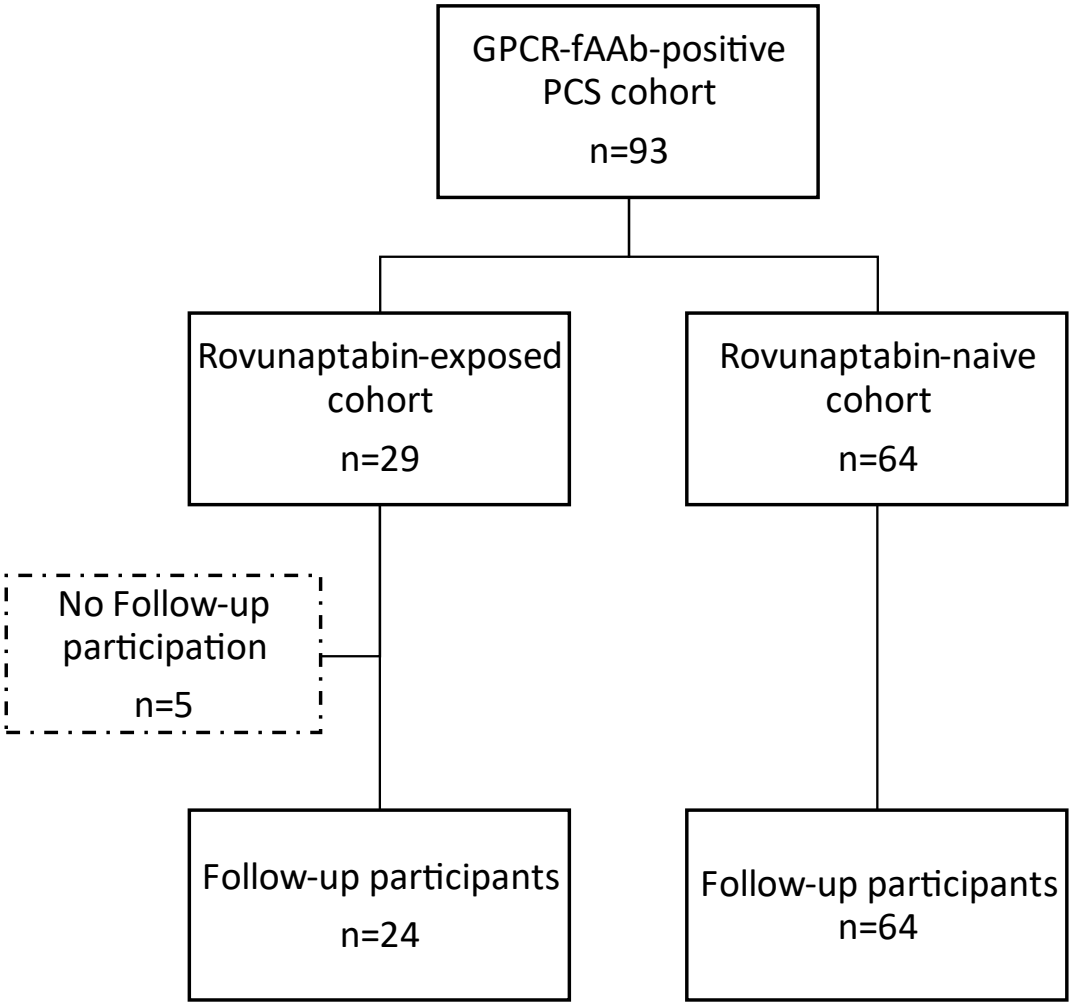
Study population and follow-up participation. Flow diagram of 93 GPCR-fAAb-seropositive patients with post-COVID syndrome (PCS), including 29 participants previously exposed to rovunaptabin (reCOVer clinical phase IIa trial) and 64 rovunaptabin-naive participants (disCOVer study). Five previously exposed participants did not participate in follow-up, resulting in 24 previously exposed and 64 rovunaptabin-naive participants included in the final analysis (n = 88).

The clinical trial reCOVer was approved by the ethics committee of the Friedrich-Alexander Universität Erlangen-Nürnberg (23-50-Az) and the German Competent Authority (Federal Institute for Drugs and Medical Devices, BfArM). The study disCOVer was approved by the ethics committee of the Friedrich-Alexander Universität Erlangen-Nürnberg (295_20 B). Written informed consent was obtained from all participants. Both studies were conducted in accordance with ICH-GCP guidelines, the Declaration of Helsinki, and national legal requirements.

### 2.2 GPCR-fAAb assessment

GPCR-fAAb seropositivity was tested using a cardiomyocyte bioassay as described previously [12, 14] for defining the breach of tolerance. Patient-derived IgG was added to neonatal rat primary cell cultures for 60 min. Depending on the direction of the Δ-frequency compared with the basal cell beating frequency, positive chronotropic GPCR-fAAbs (β2-fAAb, AT1-fAAb, α1-fAAb, Noci-fAAb) or negative chronotropic GPCR-fAAbs (MAS-AAb, M2-AAb, ETA-AAb) were detected using receptor-specific inhibitors: ICI118,551 (0.10 μM), losartan (1 μM), urapidil (1 μM), J1133977 (0.10 μM), A779 (1 μM), atropine (1 μM), and BQ123 (0.10 μM). All inhibitors were obtained from Sigma-Aldrich Chemie GmbH (Taufkirchen, Germany).

### 2.3 Patient-reported outcome measures

Patient-reported outcome measures included the Functional Assessment of Chronic Illness Therapy-Fatigue (FACIT-Fatigue) Scale 1.0, Bell score, Fatigue Severity Scale (FSS), and the 36-Item Short Form Health Survey (SF-36) [15].

The 13-item FACIT-Fatigue Scale assesses fatigue severity and its impact on functioning. Scores range from 0 to 52, with higher scores indicating less fatigue. The Bell score, with values rounded down to the nearest 10-point increment where necessary, assesses limitations in daily activities and ranges from 0 (greatest impairment) to 100 (best functioning) [16]. The nine-item FSS assesses fatigue severity and its impact on daily functioning, with higher scores indicating greater fatigue burden [17]. The SF-36 is a standardized instrument for assessing health-related quality of life. It consists of 36 items that reflect eight dimensions of physical and mental health. The individual dimensions are rated on scale from 0 to 100 points, with higher scores indicating a better health related quality of life. In addition, a physical and an mental total score can be calculated [15].

### 2.3 Statistical analysis

Continuous variables were summarized as means and standard deviations (SDs) or medians and ranges, as appropriate. Categorical variables were summarized as counts and percentages. Secondary patient-reported outcomes were analyzed descriptively using available observations at each assessment time point, without imputation of missing values. Within-participant changes in Bell score were classified as improvement, no change, or deterioration. Primary between-cohort comparison used a linear analysis of covariance (ANCOVA) model with follow-up Bell score as the dependent variable and cohort, baseline Bell score, baseline age, and sex as predictors. The model was fitted using ordinary least squares with heteroscedasticity-robust HC3 standard errors. Two-sided 95% confidence intervals (CIs) and p values were derived using the t-distribution. The cohort coefficient represents the adjusted between-cohort difference in follow-up Bell score. In exploratory analyses, Spearman rank correlations were used to examine the association between the time from the recorded positive SARS-CoV-2 test to follow-up and within-participant change in Bell score separately for each cohort. Two-sided p values were estimated using 20,000 Monte Carlo permutations with a plus-one correction, and 95% percentile CIs were calculated from 5,000 paired bootstrap resamples at the participant level. Pearson correlations served as a secondary analysis. Regression and correlation analyses were restricted to participants with complete data for the respective analysis. No adjustment for multiple comparisons was performed. Statistical analyses were conducted using R (version 4.5.1) and Python (version 3.13.6). ANCOVA and robust inference were performed using the R packages sandwich (version 3.1.3) and lmtest (version 0.9.40). Data processing, descriptive statistics, and correlation analyses were performed using pandas (version 3.1.0), NumPy (version 2.5.4), and SciPy (version 1.18.1). Figures were generated using Matplotlib (version 3.11.2).

## 3. Results

### 3.1 Nine-months follow-up: fatigue and quality of life after one-time rovunaptabin application

In the rovunaptabin exposed cohort, follow-up occurred a median of 267.5 days after baseline. Baseline data were assessed at the days directly before rovunaptabin application, respectively. Table 2 summarizes the FACIT-Fatigue, FSS, and SF-36 outcomes.

**Table 2.** Data are presented as mean (SD). Short-term follow-up assessments were obtained at visit 7 or 13, depending on the treatment sequence in the reCOVer crossover trial. Data were available for 24 participants at baseline and nine-month follow-up and for 23 participants at day 28, as one participant missed the assessment due to illness. Missing values were not imputed. Mean changes represent paired within-participant differences between baseline and nine-month follow-up (n = 24); ^1^calculated as difference between mean baseline score and the mean nine-month follow-up score.

| **Outcome** | **Baseline (n=24), Mean (SD)** | **Day 28 (n=23), Mean (SD)** | **Nine-month follow-up (n=24), Mean (SD)** | **Mean Change<sup>1</sup>** | **Changes** |
|---|---|---|---|---|---|
| **FACIT-Fatigue** | 19.83 (8.07) | 26.74 (12.61) | 29.54 (13.41) | +9.71 | Improvement |
| **FSS mean** | 6.31 (1.12) | 5.73 (1.43) | 5.29 (1.51) | −1.02 | Improvement |
| **SF-36 Vitality** | 21.67 (14.12) | 31.30 (19.55) | 38.12 (26.28) | +16.46 | Improvement |
| **SF-36 General Health** | 29.67 (16.39) | 30.96 (13.20) | 35.08 (20.55) | +5.42 | Improvement |
| **SF-36 Social Functioning** | 30.21 (24.15) | 34.78 (29.18) | 49.48 (35.64) | +19.27 | Improvement |
| **SF-36 Physical Functioning** | 46.25 (23.46) | 54.13 (26.61) | 62.50 (29.78) | +16.25 | Improvement |
| **SF-36 Role Physical** | 5.21 (20.82) | 18.48 (34.72) | 26.04 (36.47) | +20.83 | Improvement |
| **SF-36 Bodily Pain** | 40.62 (24.50) | 49.13 (25.64) | 55.42 (28.02) | +14.79 | Improvement |
| **SF-36 Physical Component Summary** | 29.52 (6.70) | 33.28 (8.69) | 41.07 (12.24) | +11.55 | Improvement |
| **SF-36 Mental Component Summary** | 36.39 (10.39) | 37.63 (11.23) | 34.89 (11.01) | −1.51 | Deterioration |

The mean FACIT-Fatigue score increased from 19.83 (SD 8.07) at baseline to 29.54 (SD 13.41) at follow-up, a mean change of +9.71 points. The mean FSS score decreased from 6.31 (SD 1.12) to 5.29 (SD 1.51), a mean change of −1.02 points. Both changes were consistent with less self-reported fatigue. The largest increases among the reported SF-36 measures were in role limitations due to physical health (+20.83 points), social functioning (+19.27), vitality (+16.46), physical functioning (+16.25), bodily pain (+14.79), and the physical component summary (+11.55). General health (+5.42) and emotional well-being (+4.00) increased by smaller amounts. The mental component summary decreased by 1.51 points.

### 3.2 Comparison of self-reported functional status between cohorts

In the previously rovunaptabin exposed cohort, the mean Bell score increased from 34.58 (SD 12.85) at baseline to 51.25 (SD 21.33) at follow-up. The mean within-participant change was +16.67 points (median +15 points). Scores increased in 17 participants (70.8%) and were unchanged in seven (29.2%). No participant reported a lower score at follow-up (Table 3, Figure 1).

**Table 3.**
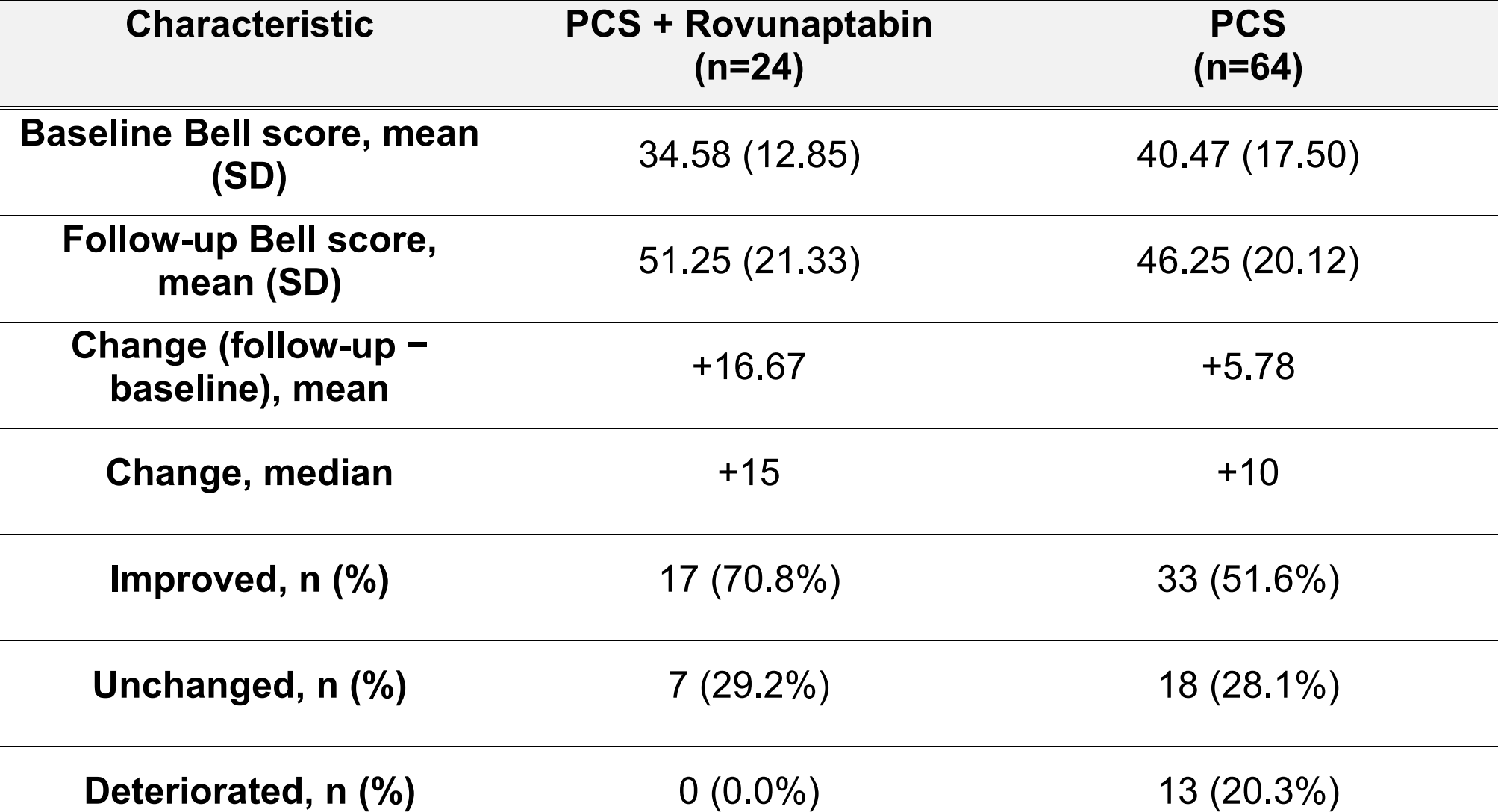
Bell score outcomes and within-participant changes from baseline to approximately nine-month follow-up in patients with post-COVID syndrome with and without prior rovunaptabin exposure. Data are presented as mean (SD), median, or number of participants (%). Improvement, no change, and deterioration were defined as an increase, no difference, or a decrease in Bell score from baseline to follow-up, respectively.

In the rovunaptabin-naive cohort, the mean Bell score increased from 40.47 (SD 17.50) at baseline to 46.25 (SD 20.12) at follow-up. The mean within-participant change was +5.78 points (median +10 points). Scores increased in 33 participants (51.6%), were unchanged in 18 (28.1%), and decreased in 13 (20.3%) (Table 3, Figure 2).

**Figure 2.**
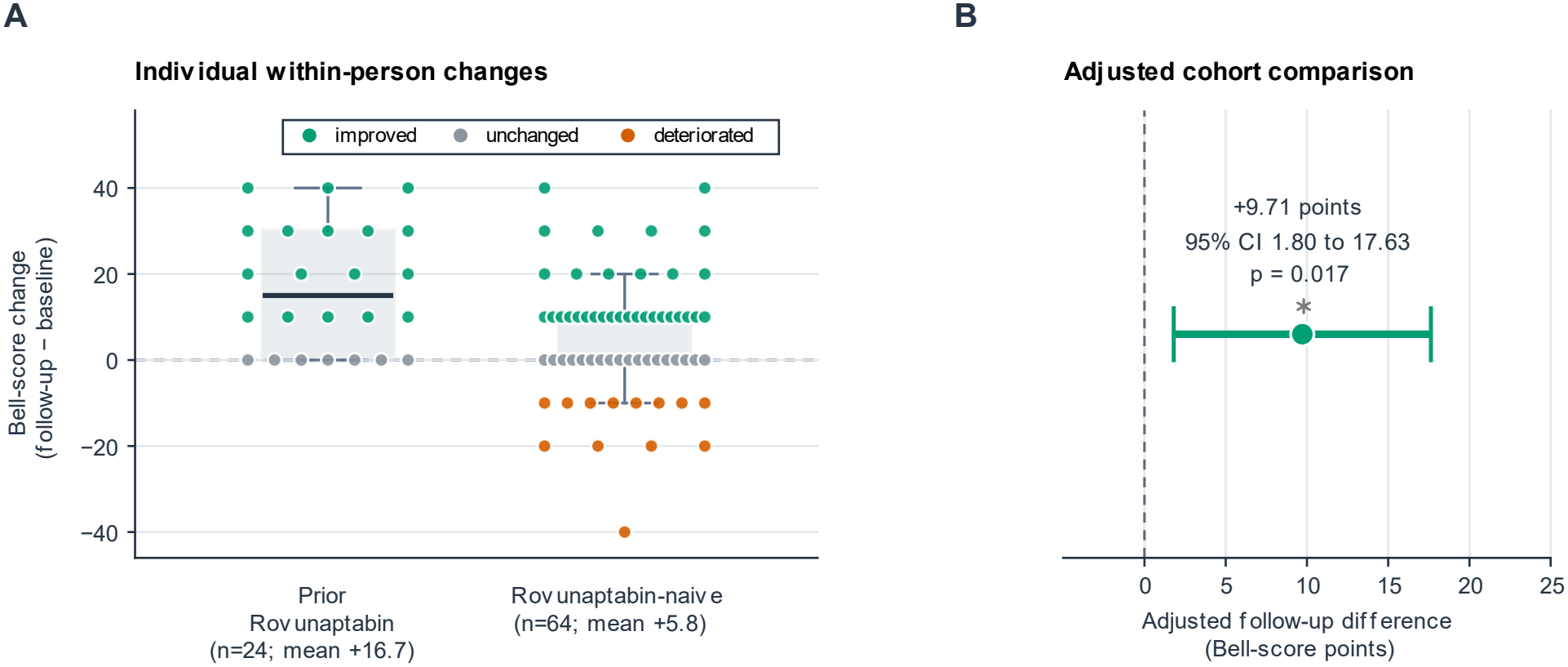
Bell score changes and adjusted follow-up difference across cohorts. (A) Individual Bell score differences (follow-up minus baseline) among participants previously exposed to rovunaptabin (n=24) and rovunaptabin-naive participants (n=64). Each point represents one participant: teal circles (improvement), gray circles (unchanged), and vermilion circles (deterioration). Boxes show median and IQR; whiskers extend to 1.5×IQR. Mean changes were +16.67 and +5.78 Bell points. (B) Between-cohort adjusted follow-up Bell score difference from ANCOVA adjusted for baseline Bell score, age and sex (HC3 robust standard errors; t-based 95% CI): +9.71 points (95% CI 1.80 to 17.63, p = 0.017).

After adjustment for baseline Bell score, age, and sex, the mean follow-up Bell score was 9.71 points higher in the previously exposed cohort than in the rovunaptabin-naive cohort (95% CI 1.80-17.63, p = 0.017). Baseline Bell score was positively associated with follow-p Be score (β 0. 3, 95 0.51-1.14, p < 0.001). ssociations with age (β 0.19, p = 0.209) and sex (β 1.84, p = 0.619) were not statistically significant.

### 3.3 Time since the positive SARS-CoV-2 test and change in Bell score

In exploratory analyses, the time from the recorded positive SARS-CoV-2 test to follow-up was not significantly correlated with Bell score change in either cohort (Figure S1).

Spearman ‘s p was −0.0 in the pre io s y exposed cohort (95 ootstrap −0. to 0.3, perm tation p 0. 25) and −0.0 in the rovunaptabin-nai e cohort (95 ootstrap −0.30 to 0.24, permutation p = 0.754). Pearson correlations were also close to zero (previously exposed cohort, r −0.091, p 0.6 3 ro napta in-nai e cohort, r −0.056, p 0.662). Consistently, the unadjusted ordinary least squares (OLS) regression lines showed small negati e s opes of −0.15 and −0.26 Be score points per month in the previously exposed and rovunaptabin-naive cohorts, respectively. These descriptive trends suggest only weak negative linear associations between time since the positive SARS-CoV-2 test and Bell score change, without evidence of statistically significant correlations.

## 4. Discussion

PCS is increasingly recognized as an umbrella syndrome encompassing multiple reproductible sub-phenotypes rather than a single homogenous disorder [18]. The subgroup framework may vary between organ-specific (e.g. pulmonary, neuropsychiatric, cardiovascular), symptom driven, severity (diminished or preserved function) or syndromic entities (e.g. myalgic encephalomyelitis/ chronic fatigue syndrome, ME/CFS, Postural Orthostatic Tachycardia Syndrome, Fibrosis). Among the proposed pathophysiological mechanisms underlying specific PCS subgroups, autoimmune processes involving GPCR-fAAbs have received increasing attention. Whether GPCR-fAAbs act as direct causal drivers of symptoms or primarily mark a distinct PCS sub-entity characterized by a breach of immunological tolerance remains an open question. In this view, GPCR-fAAb seropositivity may identify a subgroup with a specific autoimmune signature whose clinical expression, severity, and longitudinal course could differ from other PCS sub-phenotypes, rather than proving that autoantibodies alone account for all functional impairments. Several therapeutic approaches have been explored to remove or neutralize GPCR-fAAbs or target underlying immune pathways, including immunoadsorption, pharmacological neutralization, and B-cell-directed strategies [19]. Some of these modalities have been investigated in other autoantibody-associated conditions well before COVID-19; for example, immunoadsorption has been used for nearly three decades and has shown long-term clinical stability in selected cohorts such as patients with dilated cardiomyopathy [20]. However, long-term follow-up data after rovunaptabin in PCS have been lacking. The present study therefore evaluated nine-month functional outcomes in GPCR-fAAb-positive PCS patients following reCOVer trial participation compared with non-exposed seropositive patients.

The questionnaire-based follow-up aimed to evaluate patients’ perceptions and changes since the initial treatment. A significantly higher follow up Bell score in the rovunaptabin-treated cohort compared with the untreated cohort was observed nine months after application. Adjusting for baseline Bell score, age and sex, the between-group difference was 10 points (95% CI: 1.80-17.63; p = 0.017). Furthermore, baseline Bell score was associated with the follow up Bell score, showing that initial symptoms remain a factor for the long-term health status. While neither age nor sex showed a significant follow up association. In total these findings suggest that previous rovunaptabin treatment was associated with better long-term functional status, including the influence of baseline disease severity, sex and age.

The original randomized, double-blind, placebo-controlled reCOVer trial reported improvements in selected patient-reported outcomes and a reduction in GPCR-fAAb activity following rovunaptabin application [12]. The present follow-up extends the observation window and provides data on functional status beyond the original trial period. In this observational follow-up of GPCR-fAAb-positive participants, those exposed to one-time rovunaptabin reported better functional status at approximately nine months than participants who had never received rovunaptabin. The adjusted difference in follow-up Bell score was 9.71 points after accounting for baseline Bell score, age, and sex. Higher baseline Bell scores were associated with higher follow-up scores.

One proposed mechanism of rovunaptabin involves the binding and neutralization of GPCR-fAAbs. GPCRs represent a large family of membrane proteins characterized by seven transmembrane domains which play a central role in cellular communication and intracellular signaling [4]. Upon binding of an extracellular ligand, such as a hormone, neurotransmitter, or another signaling molecule, the receptor undergoes a conformational change that enable the activation of intracellular heterotrimeric G-proteins. These proteins consist of the Gα, Gβ and Gγ s nits acti ation triggers downstream signa ing pathways, inc ding the reg ation of cyclic adenosine monophosphate (cAMP), intracellular calcium levels, and protein kinase activity. To ensure controlled signaling, activated GPCRs are subject to regulatory mechanisms involving G-protein-co p ed receptor kinases (G Ks) and β-arrestin, which promote receptor desensitization and internalization [21]. Unlike conventional autoantibodies that primarily induce tissue damage, GPCR-fAAbs can also interfere with physiological processes by binding to extracellularly accessible regions of their target receptors, including extracellular loops (ECLs) and, in some cases, the extracellular N-terminal domain. This specific binding exerts their effects by modulating receptor-mediated signaling pathways [22]. Depending on their functional properties, fAAbs may act as agonists, inducing receptor activation, or as antagonists, inhibiting physiological receptor signaling [23]. Through sustained or dysregulated receptor modulation, GPCR-fAAbs may interfere with normal cellular functions and contribute to the development of pathological processes in various diseases [24]. Previous investigations have reported GPCR-fAAbs in patients with PCS and linked them to impaired microcirculation [25]. An individual treatment report described improved capillary microcirculation and fatigue symptoms after administration of rovunaptabin [12]. In the reCOVer trial, functional neutralization of GPCR-fAAb was accompanied by improvements in selected fatigue-related outcomes [13]. Viewing GPCR-fAAb seropositivity as a marker of a PCS sub-entity characterized by a breach of immunological tolerance may help explain why symptom fluctuations or renewed deterioration can occur, for instance following subsequent infections or immune triggers. However, these exploratory observations do not support routine or long-term immunosuppressive therapy in PCS, which remains unproven and requires dedicated clinical evaluation of safety and efficacy.

The original reCOVer report proposed that immunomodulatory effects involving B-cells or plasma cells might help explain symptomatic improvement beyond the immediate pharmacological activity of rovunaptabin [12]. Previous data of controlled trials show that immunoadsorption depletes autoantibodies without improving clinical symptoms, whereas observational cohorts report transient functional benefits in selected sub-phenotypes [26]. Because therapeutic apheresis does not alter the underlying autoreactive memory B-cells or prevent antibody synthesis, researchers conceptualized targeted B-cell depletion to prevent disease relapse [26]. Based on the evidence currently available, rovunaptabin has been described as an autoantibody-neutralizing DNA aptamer [6, 10]. It binds to GPCR-fAAbs and prevents them from interacting with the corresponding receptors, thereby inhibiting their receptor-activating effects [10]. Interestingly, the sustained effects observed in this study following administration of rovunaptabin may not be fully explained by its direct autoantibody-neutralizing activity alone. Although rovunaptabin has a very short plasma half-life, sustained neutralization of GPCR-fAAb has been reported following a single infusion [11]. This observation suggests that rovunaptabin may exert additional, longer-lasting immunomodulatory effects. In particular, a potential influence on B-cells and/or antibody-producing plasma cells could be involved in maintaining the reduction in circulating GPCR-fAAb levels by affecting their continued production. However, such an effect has not yet been demonstrated experimentally. The precise mechanisms underlying the sustained neutralization of GPCR-fAAb remain to be established in the future.

An important finding is the overall improvement of quality of life of the rovunaptabin exposed cohort, still after nine months after treatment with rovunaptabin. The most important finding is the significantly greater difference of Bell score changes in the rovunaptabin exposed cohort compared to the rovunaptabin-naive cohort. For the present study the Bell score was used to assess illness-related functional limitations, known to assess general functional impairment in ME/CFS [27]. In particular, it takes into account physical and mental resilience as well as the impact of the illness on daily functioning. While the FACIT Fatigue Score and the FSS enable a more nuanced assessment of fatigue symptoms, the Bell score focuses on the extend of resulting functional impairment. A key advantage of the Bell score is therefore that it captures not only subjectively perceived fatigue, but rather its functional impact on daily life. This is particularly relevant in the context of PCS, as this condition may be associated not only with fatigue but also with exercise intolerance, neurocognitive impairments, and reduced physical performance. The use of the Bell score therefore allows more comprehensive assessment of the functional burden of disease and appears particularly well-suited for evaluating limitations in daily life. Next to the improvement of fatigue, concrete improvements were observed in several SF-36 domains, including vitality, social functioning and physical functioning after nine months (Table 2). Only the Mental Component Summary showed a slight decline at the follow-up, representing a weighted total score for mental quality of life, which not only takes into account mental health, vitality, social functioning and role limitations due to emotional problems, but also incorporates contributions from the areas of physical health [28]. One possible explanation could be the SF-36’s calculation algorithm: In the classic orthogonal calculation of the Mental Component Summary, negative weighting factors are assigned to the physical dimensions. A marked improvement in physical health can therefore, from a mathematical perspective, contribute to a lower Mental Component Summary, even if individual psychological and psychosocial dimensions improve [28]. Consequently, significant improvements in physical functioning and physical pain can influence the Mental Component Summary independently of changes in individual areas of mental health. Furthermore, persistent limitations in emotional or social functioning may also have contributed to this finding [29]. Given the involvement of multiple factors, the slight decline observed should therefore be interpreted with caution and does not necessarily indicate an actual decline in mental well-being. Further analyses of the individual SF-36 domains and item-level responses would be required to clarify the underlying factors.

Importantly, baseline GPCR-fAAb seropositivity in this study served to identify an autoimmune-characterized PCS subgroup, reflecting a loss of immunological tolerance rather than a transient biomarker whose disappearance is a prerequisite for clinical improvement. Conceptually, this parallels strategies in other autoimmune-spectrum conditions - such as interventional approaches in individuals at risk for rheumatoid arthritis (e.g., the ARIAA concept) - where patients are stratified by an underlying autoimmune signature and evaluated primarily for their clinical disease course rather than complete reversal of baseline autoantibody status [30]. Whether GPCR-fAAb reactivity changes over nine months or remains detectable despite functional improvement cannot be answered from the present follow-up and should be addressed in future longitudinal studies combining clinical endpoints with serial biomarker assessments.

In both cohorts, Spearman and Pearson correlation analyses revealed weak negative, statistically non-significant associations between time since the positive SARS-CoV-2 test and changes in Bell score. The descriptive OLS trends were similarly weak. These findings provide no evidence of a significant linear association between elapsed time and Bell score improvement, although an influence of disease duration cannot be excluded. While there were improvements in the non-exposed group as well, there were also a number of cases of deterioration, unlike in rovunaptabin.

The present study is not without limitation: sample size was small and the follow-up relied on patient-reported questionnaires, which are susceptible to recall and reporting biases. Th s, f rther st dies are necessary in arger patients’ gro ps, in estigating patients se f-reported effects, morphometric and/or functional alterations as additional objective read-out. As the follow-up survey for the rovunaptabin naive group was done earlier than for the rovunaptabin exposed group (due to the later conducted reCOVer RCT), both cohorts differed markedly in time from the positive SARS-CoV-2 test to baseline.

## 5. Conclusion

This is the first study, showing 9-months long-term data after one-time application of rovunaptabin in GPCR-fAAb seropositive patients with PCS. The present data suggest that application of rovunaptabin is associated with higher follow-up Bell scores in GPCR-fAAb-positive participants with PCS. As one application of rovunaptabin showed an effect, even over a long time, larger controlled studies with longer follow-up and repeated applications of rovunaptabin are needed to assess whether (repeated) rovunaptabin produces an even enhanced lasting improvements in functional status.

## Data Availability

All data produced in the present study are available upon reasonable request to the authors.

## Contributors

**Conceptualisation:** Bettina Hohberger, Alexander Smit, Christian Mardin, Thomas Harrer, Stefanie Maas, Martin Herrmann, Marion Ganslmayer, and Jürgen Rech. **Methodology:** Bettina Hohberger, Alexander Smit, Christian Mardin, Stefanie Maas, Gerd Wallukat and Petra Lakatos. **Investigation:** Thomas Harrer, Jürgen Rech, Gerd Wallukat, Andreas Stog, Thomas Knauer, Philipp Fleischmann, Victoria Zeisberg, Adam Skornia, Monika Wytopil, Caroline Merkel, Sophia Hofmann, Julia Schottenhamml, Marion Ganslmayer, Cornelius Rosenkranz, and Jakob Hoffmanns. **Formal analysis:** Alexander Smit. **Data curation:** Alexander Smit, Bettina Hohberger, Armin Ströbel, Marion Ganslmayer, and Katja G. Schmidt. **Data access and verification:** Bettina Hohberger, Alexander Smit, Armin Ströbel, Petra Lakatos, and Katja G. Schmidt. **Visualisation:** Alexander Smit. **Validation:** Bettina Hohberger, Alexander Smit, Armin Ströbel, Petra Lakatos, and Katja G. Schmidt. **Resources:** Bettina Hohberger, Andreas Stog, Thomas Knauer, Adam Skornia, Victoria Zeisberg, Tobias Borst, Ralph Heimke-Brinck, and Alexander Bartsch. **Project administration:** Bettina Hohberger and Petra Lakatos. **Funding acquisition:** Bettina Hohberger. **Supervision:** Bettina Hohberger, Christian Mardin, and Friedrich Kruse. **Writing - original draft:** Bettina Hohberger, Alexander Smit, Thomas Knauer, Maria Reppa, Ronja Zwerenz, Valerie Hartig, Merle Flecks, Stephanie Zellinger, Philipp Fleischmann and Petra Lakatos. **Writing - review & editing:** Bettina Hohberger, Alexander Smit, Thomas Knauer, Maria Reppa, Ronja Zwerenz, Martin Herrmann, Gerd Wallukat, Valerie Hartig, Merle Flecks, Stephanie Zellinger, Philipp Fleischmann, Petra Lakatos, Cornelius Rosenkranz, Jakob Hoffmanns, Marion Ganslmayer, Friedrich Kruse, Stefanie Maas, Tobias Borst, Ralph Heimke-Brinck, Andreas Stog, Eva Rühl, Victoria Zeisberg, Adam Skornia, Alexander Bartsch, Armin Ströbel, Monika Wytopil, Caroline Merkel, Sophia Hofmann, Katja G. Schmidt, Julia Schottenhamml, Christian Mardin, Jürgen Rech, and Thomas Harrer. All authors had full access to the data and were updated at each stage of manuscript processing, including submission, revision and revision reminder, via emails from outer system or the assigned Assistant Editor.

## Data sharing

The dataset analyzed is not publicly available due to general data privacy regulations, but can be requested from the corresponding author upon reasonable request.

## Acknowledgements

The study was funded by the Bavarian State Ministry of Science and Arts (StMWK, Bavarian Health and Food Safety Authority (LGL), 2490-PC-2021-V14, Discover-Establishment and evaluation of clinical algorithms for objective diagnosis of subtypes of Long-COVID as an essential basis for effective patient care), the German Research Foundation (DFG; GRK 2504/1, Novel antiviral approaches from small molecules to immune intervention; 401821119/GRK2504), and the German Federal Ministry for Research, Technology and Space (BMFTR; 01EO2105, iIMMUNE_ACS-Förderung von Advanced Clinician Scientists im Bereich Immunmedizin; 01EP2108A, reCOVer). The funders had no role in the study design, conduct, data collection, analysis and interpretation, or preparation of the manuscript. The Department of ophthalmology is part of the Universität of Erlangen-Nürnberg, Friedrich Alexander-Universität Erlangen-Nürnberg (FAU), Germany. AI tools were used to check grammar and word choice in the writing of the report, since we are not native to the English language. AI was not used for analysis.

## Supplementary material

**Figure S1.**
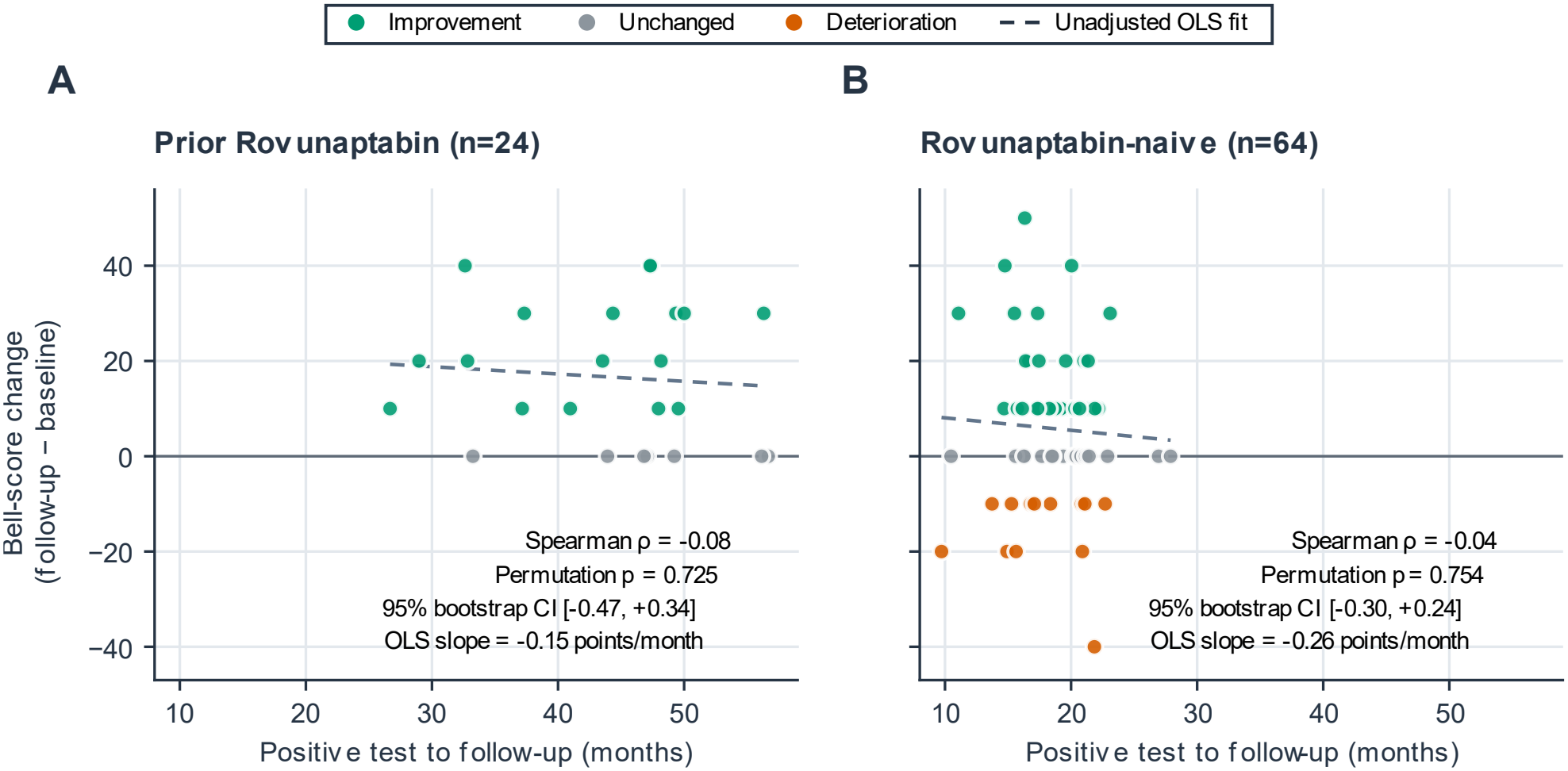
Time from positive SARS-CoV-2 test to follow-up vs. within-person Bell score change in previously exposed (A; n=24) and naive (B; n=64) cohorts. Markers: teal circles (improvement), gray circles (unchanged), and vermilion circles (deterioration). Time in months equals elapsed days divided by 30.4375. Dashed cohort-specific lines are descriptive unadjusted ordinary least square (OLS) trends. OLS slopes: −0.15 and −0.26 points/month. Spearman rho with two-sided permutation p-values (20,000 permutations) and paired bootstrap 95% percentile CIs (5,000 resamples) are displayed within each panel. The exploratory analyses are unadjusted; a nonsignificant association does not establish independence from disease duration.

